# A Blood Based Mitochondrial Functional Index Biomarker for Alzheimer’s Disease

**DOI:** 10.1101/2025.09.05.25335197

**Authors:** Brittany M. Hauger, Riley E. Kemna, Paul J. Kueck, Taylor A. Strope, Casey S. John, Keith P. Smith, Hana D. Mayfield, Ellen Herrold, Keri L. Cox, Rebecca Bothwell, Leonidas Bantis, Russell H. Swerdlow, Jill K. Morris, Heather M. Wilkins

## Abstract

**INTRODUCTION;:** Alzheimer’s disease (AD) pathology is complex and involves mitochondrial dysfunction. There are emerging therapies targeting mitochondrial function in clinical trials for AD. This highlights the need for biomarkers that measure mitochondrial function.

**METHODS;:** We determined the utility of a novel blood based mitochondrial biomarker, the mitochondrial functional index (MFI) in the context of AD.

**RESULTS;:** *In vitro* and *in vivo* models of AD had a reduced MFI. MFI was lower in human AD subjects and *APOE4* carriers. ROC analysis for individual biomarkers was completed and MFI had a higher area under the curve than other plasma biomarkers. The MFI biomarker correlated with the Mini Mental State Exam (MMSE) and the clinical dementia rating scale (CDR).

**DISCUSSION;:** This study highlights the potential utility of MFI as a functional blood based mitochondrial biomarker to interrogate energy metabolism. Ongoing studies are examining the relationship of MFI with brain energy metabolism outcomes.

## Background

Alzheimer’s disease (AD) pathology begins decades before clinical onset of dementia. Amyloid beta (Aβ) generally accumulates first in cognitively normal individuals, with tau and cognitive abnormalities following ^1^. Metabolic changes are prominent in AD and mitochondrial function correlates with amyloid and tau measures in cell and animal AD models in addition to human subjects with AD^2–7^.

Fluorodeoxyglucose positron emission tomography (FDG-PET) comparing AD and cognitively normal individuals reveals lower glucose uptake or metabolism in the brain of AD patients ^8–11^. Beyond reductions in brain glucose metabolism, mitochondrial dysfunction is observed not only within the brain but also systemically in AD ^12–21^. More recent genome wide association studies (GWAS) identified risk-associated single nucleotide polymorphisms (SNPs) in genes which function in mitochondrial and metabolic pathways ^22–25^. Apolipoprotein E (*APOE*), the strongest genetic risk factor for sporadic AD, is central to lipid metabolism and has been found to interact with inherited mitochondrial genes to amplify risk for AD ^25–27^. Moreover, postmortem AD brain has an overall reduction in the number of intact mitochondria and mitochondrial DNA (mtDNA) ^21, 28^. Mitochondrial dysfunction plays a role in protein aggregation, inflammation, and cell death--all events observed in AD. Overall, metabolism and mitochondrial dysfunction are strongly associated with AD.

Targeting metabolic and mitochondrial function in clinical trials is an emerging theme for AD. This highlights the importance and need for biomarkers which measure mitochondrial function in a non-invasive manner. Overall AD lacks adequate available therapies. Drug and lifestyle intervention development have been difficult due to a lack of therapeutic response biomarkers. Biomarkers are essential for therapeutic response outcomes for AD and to facilitate treatment development efforts.

Measuring mitochondrial function in blood could serve as a powerful tool to understand mitochondrial dynamics in disease and as potential response biomarkers for clinical trials. However, mitochondrial function measures can be complex, noisy, and unreliable. We have published using prior blood based mitochondrial measures in therapeutic clinical trials for ALS and AD as secondary response outcomes^29–35^. Some of these mitochondrial measures were flow cytometry based fluorescent measures of mitochondrial superoxide, mitochondrial membrane potential, and apoptosis. These individual measures had high variability, so we worked to develop an algorithm to overcome these issues and provide a comprehensive measure to assess mitochondrial function. Here, we describe a novel blood based mitochondrial biomarker that is measured from peripheral blood mononuclear cells (PBMCs) using fluorophores and flow cytometry. Measures are used in a log-based algorithm to calculate a Mitochondrial Functional Index (MFI). Here we describe the potential utility of the MFI biomarker in AD.

## Methods

### Human Subjects

This study was approved by the Ethics Committee at the University of Kansas Medical Center (FWA00003411). All participants provided written informed consent prior to enrolment in the study. This research was conducted ethically in accordance with the World Medical Association Declaration of Helsinki. We enrolled 20 non-demented (ND), 20 mild cognitive impairment (MCI), and 19 AD subjects from the University of Kansas Alzheimer’s Disease Research Center (KU ADRC) clinical cohort. Inclusion criteria for this group included individuals diagnosed with Alzheimer’s Disease, Mild Cognitive Impairment, or probable Alzheimer’s Disease, or cognitively normal individuals, at least 18 years of age, and patients who were willing to give informed consent. Exclusion criteria included diagnosis of other neurodegenerative diseases (Parkinson disease, ALS, etc), clinically significant history of unstable medical illness (unstable angina, advanced cancer, etc) over the last 30 days, limited mental capacity such that the patient cannot provide written informed consent or comply with evaluation procedures, history of recent alcohol or drug abuse or noncompliance with treatment or other experimental protocols. Mini mental state exam (MMSE) and clinical diagnostic rating (CDR) data were collected from the KU ADRC cohort. These exams are administered to clinical cohort participants yearly.

We also enrolled 10 healthy volunteers with the following inclusion criteria; no diagnosis of neurodegenerative disease or other significant clinical illness, between 18 and 35 years old, and patients who were willing to give informed consent. Exclusion criteria included diagnosis of neurodegenerative diseases (AD, dementia, Parkinson disease, ALS, etc), clinically significant history of unstable medical illness (unstable angina, advanced cancer, etc) over the last 30 days, history of recent alcohol or drug abuse or noncompliance with treatment or other experimental protocols, persons with excessively low blood pressure (systolic blood pressure <85, diastolic blood pressure <55), excessive bradycardia (pulse <55), or tachycardia (pulse >100), persons who have had adverse events such as fainting or excessive bleeding during a past phlebotomy, persons who had a greater than 100 cc phlebotomy within the past 2 weeks prior to the study, or who are anticipated to undergo more than 100 cc of phlebotomy during the next two weeks of the study. Human subject demographics for ND, AD, and MCI groups are represented in **Table 1**. Healthy volunteers had an average age of 28.6 years with a standard deviation of 2.7 years, all white/non-Hispanic, 5 male and 5 female subjects.

**Table 1.**
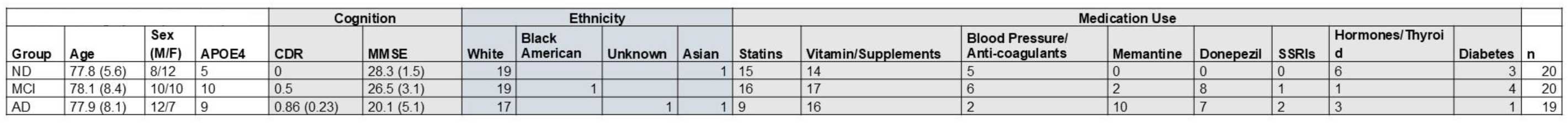
Demographics (Mean SD)

### Study Visits

For ND, MCI, and AD subjects only one study visit was completed. After subjects were consented a phlebotomy was performed were three ACD tubes (8.5 mL each) and one EDTA tube (9 mL) for a total phlebotomy volume of ∼35 mL was collected.

For healthy volunteers, the study involved four visits. For visit one, after informed consent, subjects received a questionnaire asking about medications/supplement use, alcohol use, and smoking history. Subjects underwent a fasting phlebotomy in three ACD tubes (8.5 mL each) and one EDTA tube (10 mL) for a total volume of ∼36 mL. For visit two subjects underwent a non-fasting phlebotomy of three ACD tubes (8.5 mL each) and one EDTA tube (10 mL) for a total volume of ∼36 mL. Subjects completed a dietary recall questionnaire. For visit three subjects underwent a fasting phlebotomy in three ACD tubes (8.5 mL each), three EDTA tubes (9 mL each), and three heparin tubes (10 mL each) for a total volume of ∼83 mL. For visit four subjects underwent a fasting phlebotomy in twelve ACD (8.5 mL each) tubes for a total volume of ∼102 mL.

### Blood Processing

PBMCs were isolated using Accuspin tubes, histopaque 1077, and differential centrifugation as previously described ^30, 31, 34^. For mitochondrial biomarkers in the ND, MCI, and AD groups blood was collected in ACD tubes and processed/measured within 30 hours of blood draw. For visits one, two, and three of the healthy control group blood was collected in varying anticoagulant types (EDTA, ACD, and heparin) and blood was processed/measured for mitochondrial biomarkers the same day. For visit four of the healthy control group blood was collected in ACD tubes and processed/measured for mitochondrial biomarkers over varying times (less than 24 hours, 24 hours, 48 hours, 72 hours, and 96 hours) following phlebotomy. For A/T/N analyses blood was collected in EDTA tubes and spun down at 500 x g for 15 minutes. Plasma was collected, aliquoted, and frozen until measures were completed.

### iPSC Source and Reprogramming

iPSCs were reprogrammed from dura fibroblasts obtained at the KU ADRC or purchased from WiCell. KU ADRC fibroblast donors were members of the Clinical Cohort, who consented to donation upon death and approval from an ethical standards committee to conduct this study was received. The studies involving human participants were reviewed and approved by the University of Kansas Medical Center Institutional Review Board. Banked tissue is de-identified by the KU ADRC Neuropathology Core to eliminate identifying information. Reprogramming was completed using the Sendai Virus, CytoTune-iPS 2.0 *Sendai Reprogramming* Kit from ThermoFisher. iPSC were age, sex, and diagnosis matched (n=4 per diagnosis). For iPSCs derived from the KU ADRC cohort, ND or AD were diagnosed at autopsy neuropathological examination as outlined in the NACC Neuropathology Coding Guidebook^36^.

### iPSC neural progenitor cell differentiation

iPSCs were differentiated into neural progenitor cells (NPCs) using STEMDiff Neural Induction Medium (NIM). iPSCs were placed into a single cell suspension in NIM with SMADi/ROCKi (SMAD inhibitor and ROCK Inhibitor) in an AggreWell 800 plate. Embryoid bodies were cultured in the AggreWell plate for 5 days with NIM/SMADi partial medium changes daily. Embryoid bodies were then plated onto Matrigel coated plates and fed daily with NIM/SMADi medium until day 12 to allow neural rosette formation. Neural rosettes were selected using Neural Rosette Selection Reagent (StemCell Tech) and plated onto Matrigel coated dishes with NIM/SMADi. Media was changed daily for 7 days, after which neural progenitor cells (NPC) were cryopreserved and split into defined Neural Progenitor Medium (StemCell Tech).

### iPSC forebrain neuronal differentiation

NPCs were plated onto PLO/Laminin coated dishes in neural progenitor medium. The following day media was changed to StemDiff Forebrain Neural Differentiation Medium (StemCell Tech). Media was changed daily for 7 days ^37, 38^. Following which cells were plated onto PLO/laminin coated dishes in defined Brain Phys Medium (with N2A, SM1, BDNF, GDNF, cAMP, and ascorbic acid) for neuronal maturation. Neurons were matured for 7-10 days and used for downstream experiments.

### iPSC astrocyte differentiation

NPCs were plated onto Matrigel coated dishes in neural progenitor medium. The following day cells were placed in astrocyte differentiation medium consisting of DMEM (high glucose, with glutamine, no pyruvate), B27, 1% FBS, glutamine, bFGF, CNTF, BMP8, Activin A, heregulin 1b, and IGF1. Medium was changed every other day and cells were passaged as needed ^39^. After 30 days or approximately 5-6 passages astrocytes were used in downstream experiments.

### 5xFAD mice

All animals were housed in a temperature (∼25°C) and light-controlled (12:12 hour light-dark) room with free access to food and water. Mice were euthanized in the morning between 8:00-10:00 a.m., with CO_2_ and decapitation, where trunk blood was collected in ACD solution. PBMCs were isolated using histopaque 1077 and differential centrifugation. The Institutional Animal Care and Use Committee (A3237-01) approved the animal protocols at the University of Kansas Medical Center. n= 3 per group (2 males, 1 female) at ∼8 weeks of age.

### Mitochondrial Functional Index

Approximately 2 million PBMCs were stained with Annexin V+MitoTracker, MitoSox+ Hoechst, and TMRE+ Hoechst as previously described ^30, 31^. Briefly, cells were incubated with 40 nM MitoTracker, 5 µM MitoSox/10 ng Hoechst, and 200 nM TMRE/10 ng Hoechst (in separate tubes) for 30 minutes at 37°C/5% CO_2_ in Hanks Balanced Salt Solution (HBSS with Ca2+/Mg2+). MitoTracker cells are washed and then stained with Annexin V in binding buffer, then diluted for flow cytometry analysis. MitoSox and TMRE stained cells are washed with HBSS and diluted for flow cytometry analysis, where 10,000 cells per tube were analyzed. All values were normalized to Hoechst signal. Fluorescent measures were completed within 30 hours of blood draw. The MFI algorithm; MFI = log [(MitoTracker x TMRE)/(MitoSox x Annexin V)] provides an overall picture of mitochondrial health and function. The MFI biomarker is listed in utility patent no. 63/824,391 *(Mitochondrial Functional Index)* as of June 16, 2025.

### A/T/N Measures

Whole blood was collected via venipuncture into vacutainer tubes containing EDTA as an anticoagulant. Blood was centrifuged at 1800 x g for 10 minutes to separate plasma, which was aliquoted and stored at –80C prior to analyses. Fluid biomarkers were analyzed on Quanterix Simoa HD-X (Neuro 4 Plex E (N4PE; Aβ40, Aβ42, NfL, GFAP) and pTau181 v2.0 and 2.1 Advantage) per manufacturer instructions using appropriate calibrators and controls. Because samples were analyzed using two versions of the pTau181 assay, a conversion factor published by Quanterix was employed to adjust values for samples analyzed on pTau181 V2.1.

### APOE genotyping

A single nucleotide polymorphism (SNP) allelic discrimination assay was used to determine APOE genotypes. This involved adding 5 ul of blood to a Taqman Sample-to-SNP kit (ThermoFisher). Taqman probes to the two APOE-defining SNPs, rs429358 (C_3084793_20) and rs7412 (C-_904973_10) (ThermoFisher), were used to identify APOE ε2, *ε3*, and *ε4* alleles.

### Data Analysis

All experiments were completed at least three different sets of differentiated cells or mice. Data were summarized by means and standard deviations or errors. To compare means between three or more groups, we used one-way ANOVA followed by Fisher’s least significant difference (LSD) post hoc testing. To compare means between two groups we used two-way, unpaired Student’s t-tests. Receiver Operating Characteristic (ROC) analysis was completed using the Wilson Brown Method. Correlation analyses used the Pearson’s test. Statistical tests were performed using Prism/Graph pad. p-values less than 0.05 were considered statistically significant.

## Results

The mitochondrial functional index (MFI) biomarker combines mitochondrial reporter dyes for mitochondrial membrane potential (TMRE), mitochondrial superoxide (MitoSox), mitochondrial mass (MitoTracker), and apoptosis (Annexin V) in an algorithm to assess overall mitochondrial function. A higher MFI value is associated with higher mitochondrial function. In human iPSC derived neurons from a healthy donor, treatment with rotenone and antimycin A significantly reduced the MFI biomarker (**Figure 1A**). Rotenone and antimycin A are complex I and complex III inhibitors, respectively. We next examined the levels of the MFI biomarker in induced pluripotent stem cell (iPSC) derived neurons and astrocytes from ND and sporadic AD subjects. The MFI biomarker was significantly reduced in both neurons and astrocytes derived from AD subject iPSCs (**Figure 1B,C**). To further examine if the MFI biomarker is altered in models of AD, we examined it in PBMCs from 5xFAD mice at 8 weeks of age. There was a significant reduction in MFI in the 5xFAD mice (**Figure 1D**).

**Figure 1.**
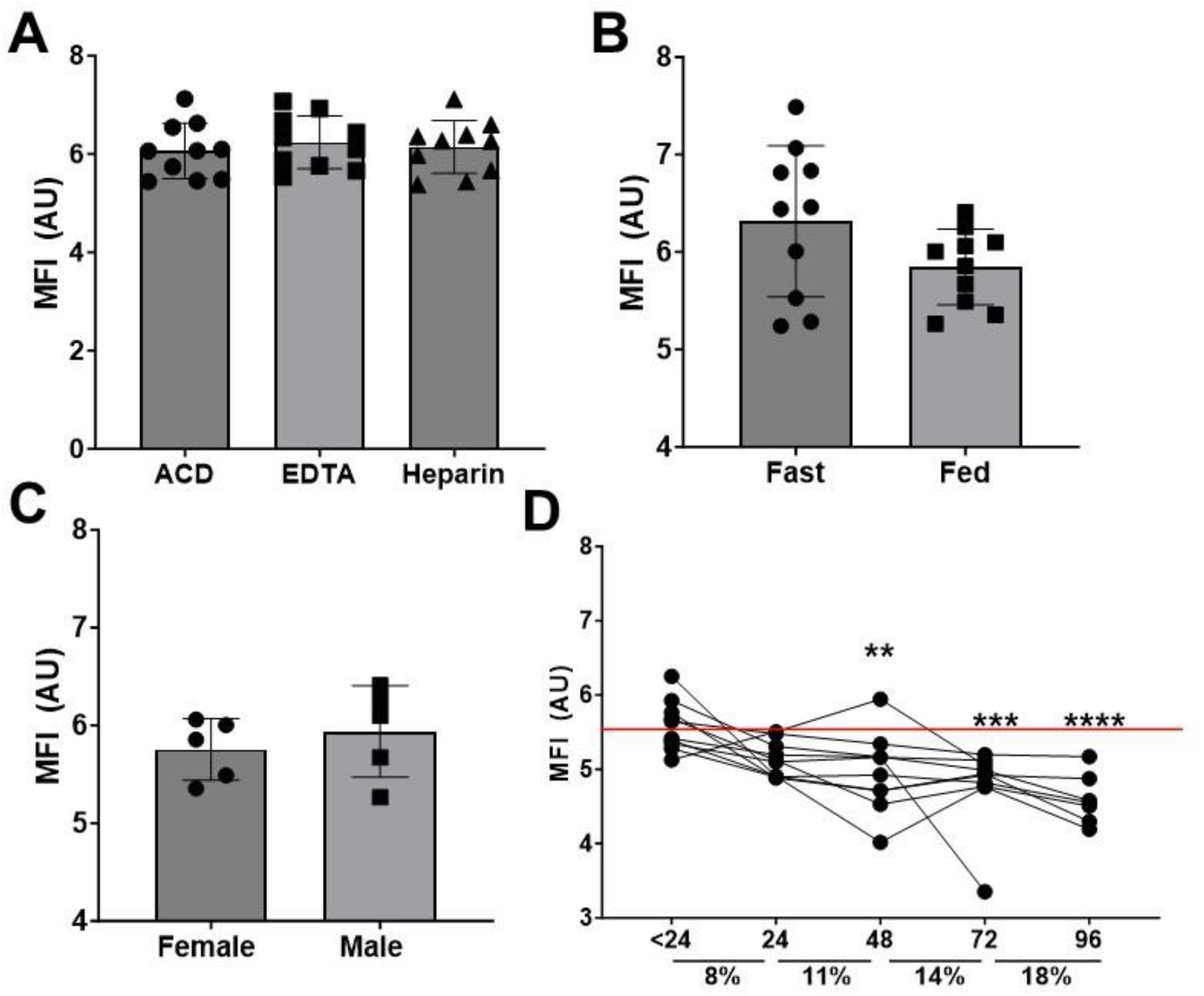
MFI Biomarker Development. A. MFI measures with different anticoagulant types including ACD, EDTA, and Heparin. n=10. B. MFI in fasting versus fed state, n=10. C. MFI by sex, n=5 each. D. MFI following different blood processing times y axis indicates hours since blood draw and % reduction in biomarker following the time point prior. Mean with SD.

To establish standardized protocols and stability for the MFI biomarker in human blood, we enrolled 10 healthy (18-35 years old, 5 male/5 female) individuals. No difference in MFI biomarker levels was observed between ACD, EDTA, or Heparin anticoagulants (**Figure 2A**). We observed no difference in MFI biomarker level between fasting or fed blood draws (**Figure 2B**) or between male and female participants (**Figure 2C**). The stability of the MFI biomarker showed significant reductions in levels between 24- and 48-hours following blood collection in ACD tubes (**Figure 2D**). With each 24-hour period there was an approximate 8-18% reduction in MFI biomarker levels. Based on these findings all biomarker processing was completed within 30 hours of blood collection for this study.

**Figure 2.**
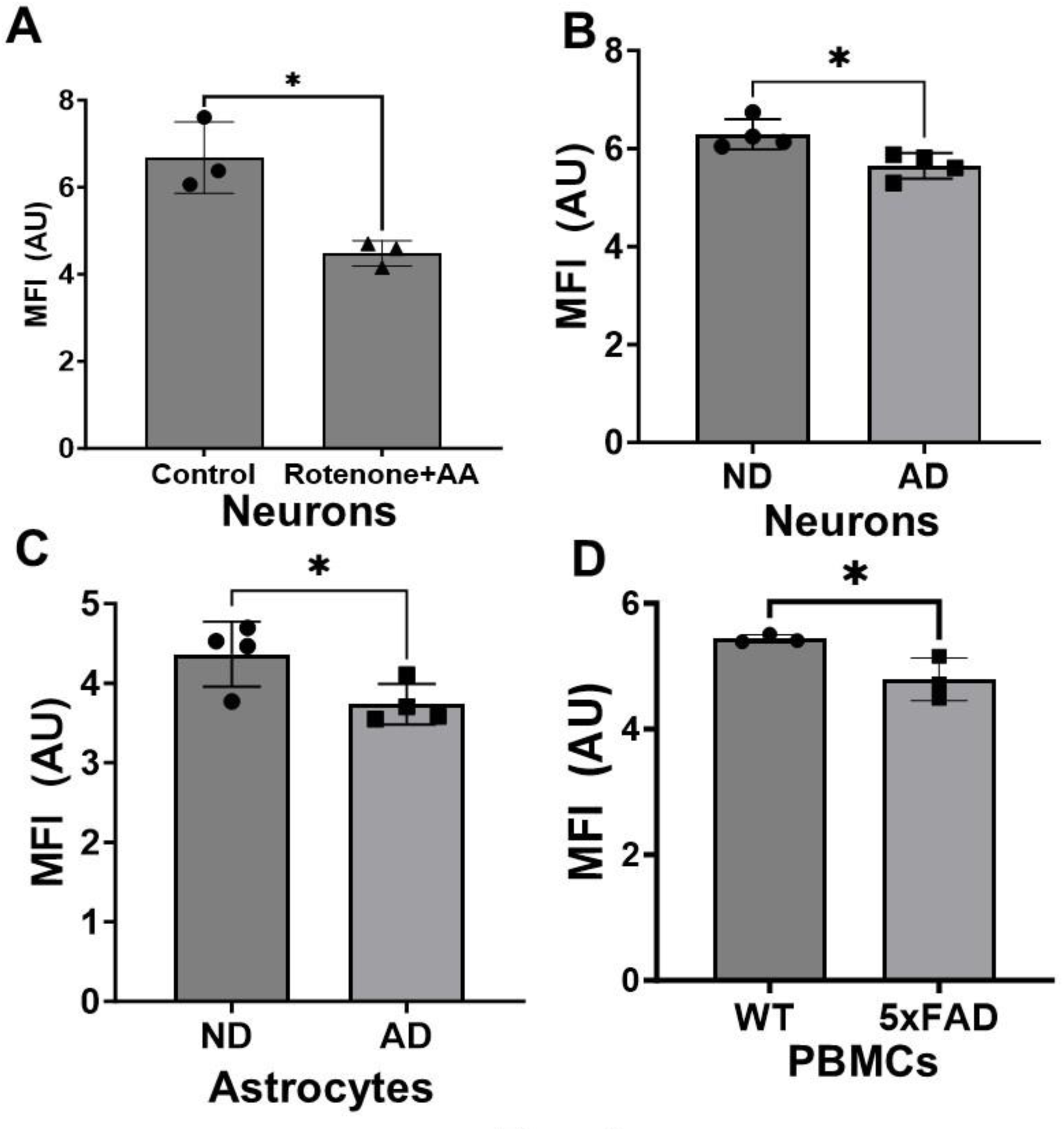
MFI Biomarker in AD models. A. MFI in iPSC derived neurons treated with mitochondrial toxins (1 µM each rotenone and antimycin A, 1 hour), n=3 per group. B. MFI measure in iPSC derived neurons, n=4 per group. C. MFI measure in iPSC derived astrocytes, n=4 per group. D. MFI measure in 5xFAD mice (2 males, 1 female per group) at age 8 weeks from PBMCs, n=3 per group. Mean with SD.

We next examined the MFI biomarker in ND, MCI, and AD subjects. A significant reduction in MFI biomarker was observed in the AD group when compared to the ND group (**Figure 3A**). The MFI biomarker was also lower in individuals that carry an *APOE4* allele regardless of diagnosis (**Figure 3B**). When examining ROC analysis of the MFI biomarker it showed an AUC of 0.8028 between ND and AD, 0.6025 between ND and MCI, and 0.8139 between MCI and AD (**Table 2**, **Figure 3C**). The MFI is composed of four biomarkers in its algorithm and each individual biomarker AUC for discrimination between ND and AD are shown in **Table 3**. The AUC ranged from 0.5526-0.622 for individual biomarkers compared to 0.8028 for the MFI algorithm. Plasma A/T/N biomarkers showed significant differences between ND and AD groups for pTau181, NfL, and GFAP (**Table 4**). The MCI group had significantly increased plasma pTau181 values compared to the ND group (**Table 4**). When comparing the MFI biomarker performance to other plasma ATN biomarkers, the MFI showed a higher AUC for discrimination between ND and AD, and MCI and AD over pTau181, GFAP, NfL, and Aβ_42/40_ (**Table 5**).

**Figure 3.**
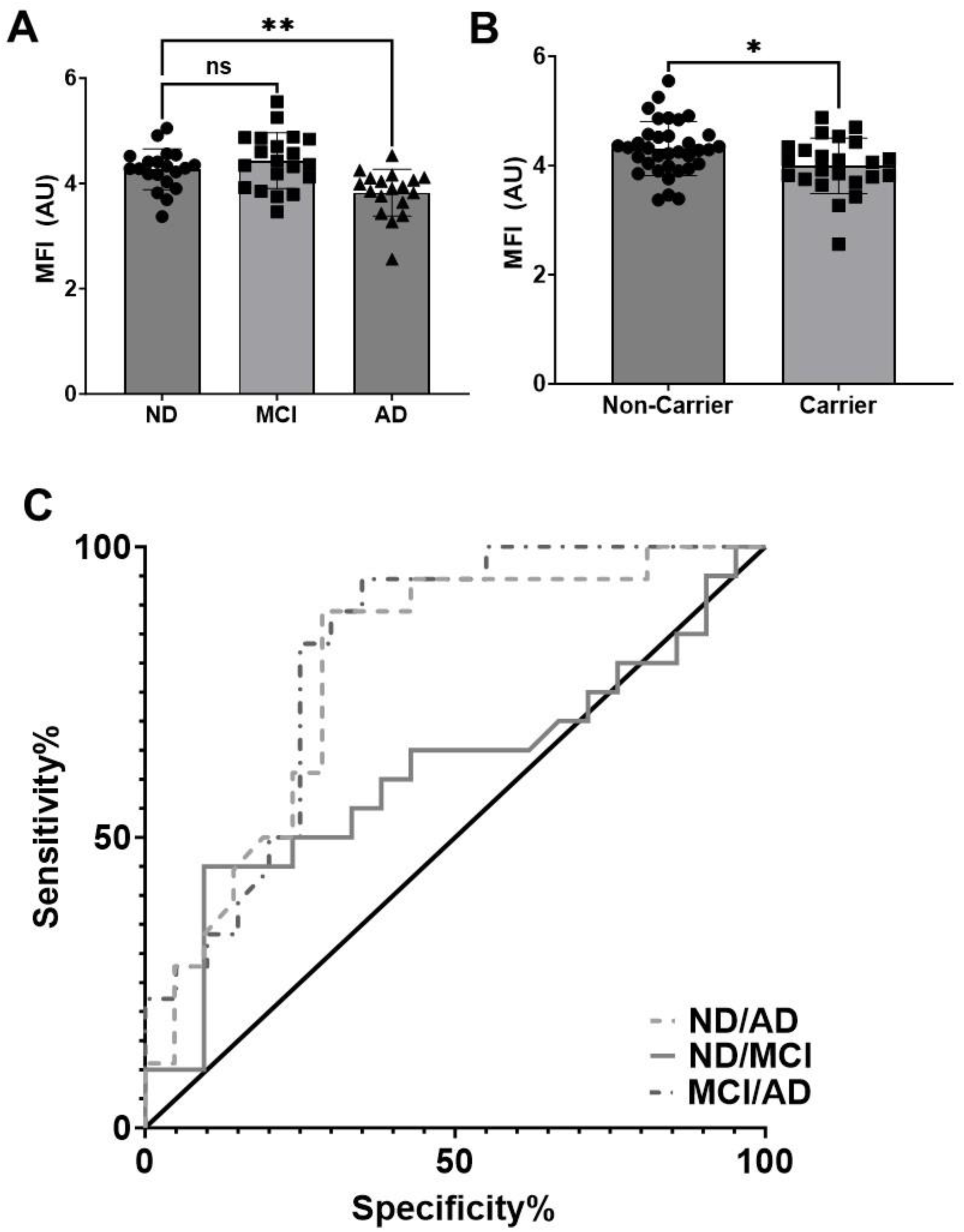
MFI Biomarker in AD. A. MFI measures in ND, MCI, and AD. n=20 for ND and MCI, n=19 for AD. B. MFI by *APOE4* carrier status n=34 non-carriers, n=23 carriers. C. AUC analysis discrimination between ND/AD, ND/MCI, or MCI/AD. Mean with SD.

**Table 2.** MFI Biomarker ROC.

| Table 2. MFI Biomarker ROC | AUC | SE | CI_lower | CI_upper | p-value | Sens at 95% Spec | Spec at 95% Sens |
| --- | --- | --- | --- | --- | --- | --- | --- |
| MFI (ND v AD) | 0.8028 | 0.0729 | 0.6599 | 0.9457 | 0 | 0.2 | 0.1111 |
| MFI (ND v MCI) | 0.6025 | 0.0932 | 0.4198 | 0.7852 | 0.1358 | 0.1 | 0.05 |
| MFI (MCI v AD) | 0.8139 | 0.069 | 0.6787 | 0.9491 | 0 | 0.45 | 0.2222 |

**Table 3.** Individual Biomarker ROC ND v AD.

| Table 3. Individual Biomarker ROC<br>ND v AD | AUC | SE | CI_lower | CI_upper | p-value | Sens at 95% Spec | Spec at 95% Sens |
| --- | --- | --- | --- | --- | --- | --- | --- |
| TMRE | 0.6222 | 0.0938 | 0.4384 | 0.8061 | 0.0963 | 0.05 | 0.1111 |
| MitoSox | 0.5658 | 0.094 | 0.3816 | 0.75 | 0.242 | 0.1053 | 0.05 |
| Annexin V | 0.5961 | 0.0895 | 0.4206 | 0.7715 | 0.1417 | 0.1053 | 0.05 |
| MitoTracker | 0.5526 | 0.0943 | 0.3678 | 0.7375 | 0.2884 | 0.1 | 0 |

**Table 4.** Plasma Biomarkers (Mean SD)

| Table 4. Plasma Biomarkers (Mean SD) |  |  |  |  |
| --- | --- | --- | --- | --- |
| Group | Aβ<sub>42</sub>/Aβ<sub>40</sub> | pTau181 | GFAP | NfL |
| ND | 0.089 (0.014) | 2.42 (0.91) | 158.2 (66) | 18 (6.5) |
| MCI | 0.0819 (0.0168) | **3.89 (2)\*** | 170.4 (80) | 18.8 (7.4) |
| AD | 0.0847 (0.0117) | **3.80 (1.5)\*** | **228.9 (101)\*** | **24.1 (8.9)\*** |
| ANOVA | 0.2076 | **0.0085** | **0.0277** | **0.0375** |

**Table 5.** Plasma Biomarker Compared to MFI Biomarker ROC Analysis.

| Table 5. Plasma Biomarker Compared to MFI Biomarker ROC Analysis |  |  |  |  |  |  |  |  |
| --- | --- | --- | --- | --- | --- | --- | --- | --- |
| ND v AD | Biomarker | AUC | SE | CI_lower | CI_upper | p-value-1t | Sens at 95% Spec | Spec at 95% Sens |
|  | Ptau181 | 0.7796 | 0.08 | 0.62 | 0.94 | 0.00 | 0.50 | 0.11 |
|  | GFAP | 0.7194 | 0.08 | 0.56 | 0.88 | 0.00 | 0.39 | 0.05 |
|  | NfL | 0.7194 | 0.08 | 0.56 | 0.88 | 0.00 | 0.28 | 0.10 |
| | Aβ<sub>42</sub>/Aβ<sub>40</sub> | 0.6658 | 0.09 | 0.49 | 0.84 | 0.03 | 0.00 | 0.00 |
|  | MFI | 0.8028 | 0.07 | 0.66 | 0.95 | 0.00 | 0.20 | 0.11 |
| ND v MCI | Biomarker | AUC | SE | CI_lower | CI_upper | p-value-1t | Sens at 95% Spec | Spec at 95% Sens |
|  | Ptau181 | 0.7105 | 0.09 | 0.53 | 0.89 | 0.01 | 0.44 | 0.00 |
|  | GFAP | 0.5737 | 0.10 | 0.38 | 0.76 | 0.22 | 0.21 | 0.00 |
|  | NfL | 0.5368 | 0.10 | 0.35 | 0.73 | 0.35 | 0.21 | 0.00 |
| | Aβ<sub>42</sub>/Aβ<sub>40</sub> | 0.6700 | 0.09 | 0.50 | 0.84 | 0.03 | 0.05 | 0.05 |
|  | MFI | 0.6025 | 0.09 | 0.42 | 0.79 | 0.14 | 0.10 | 0.05 |
| MCI v AD | Biomarker | AUC | SE | CI_lower | CI_upper | p-value-1t | Sens at 95% Spec | Spec at 95% Sens |
|  | pTau181 | 0.5078 | 0.11 | 0.30 | 0.72 | 0.47 | 0.00 | 0.13 |
|  | GFAP | 0.5965 | 0.10 | 0.40 | 0.79 | 0.16 | 0.00 | 0.11 |
|  | NfL | 0.6462 | 0.09 | 0.46 | 0.83 | 0.06 | 0.00 | 0.05 |
| | Aβ<sub>42</sub>/Aβ<sub>40</sub> | 0.5632 | 0.10 | 0.38 | 0.75 | 0.25 | 0.05 | 0.15 |
|  | MFI | 0.8139 | 0.07 | 0.68 | 0.95 | 0.00 | 0.45 | 0.22 |

The MFI biomarker significantly correlated with MMSE and CDR (**Figure 4**). MMSE showed significant correlation with CDR as well. Plasma NfL correlated with age, plasma GFAP, and plasma pTau181 (**Figure 4**).

**Figure 4.**
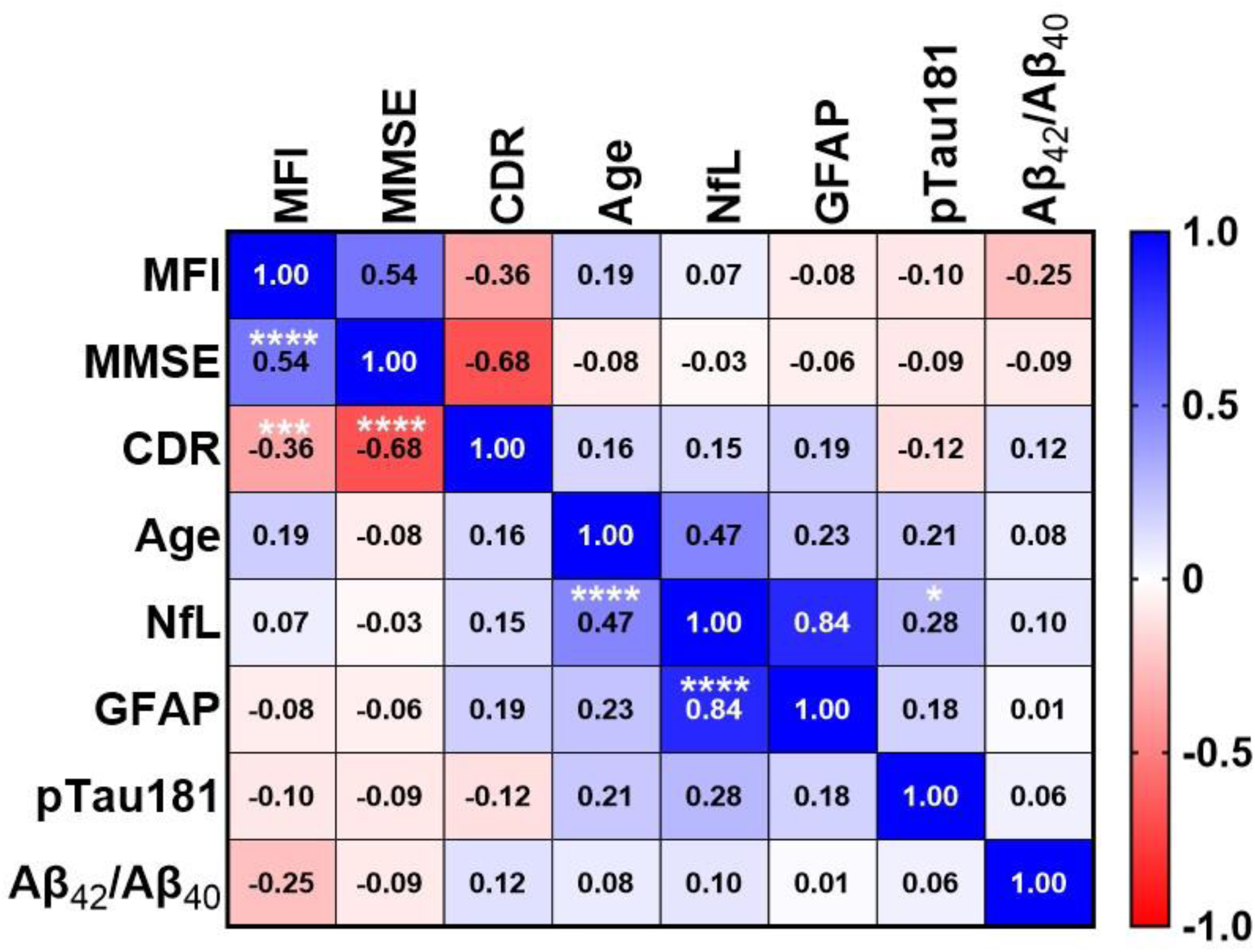
MFI correlation with other biomarkers. Correlation matrix between MFI and all other biomarkers, numbers show Pearson’s R. Significant correlations were observed between: MMSE/MFI p=0.00015, CDR/MFI p=0.005, pTau181/NfL p=0.05, Age/NfLp=0.00019, GFAP/NfL p=1.73×10^−16^.

Finally, we examined if commonly used therapies for AD that have been implicated in modulating mitochondrial function affect the MFI biomarker. We saw no evidence that donepezil affected the MFI biomarker in the AD group (data not shown).

## Discussion

Current blood-based biomarkers for AD focus on amyloid, tau, and neurodegeneration (A/T/N) measures in plasma. Amyloid and tau levels are relatively straightforward, however measuring neurodegeneration in plasma utilizes surrogates of neuronal and glial brain status, which include NfL and GFAP. Prior studies have reported that those with AD have elevated plasma phosphorylated tau (pTau181 or pTau217), reduced Aβ_42_/Aβ_40_ ratios, increased NfL, and increased GFAP ^40–46^. The MFI biomarker significantly correlated with NfL but no other plasma biomarkers measured in this study. As expected GFAP and NfL correlated as well as NfL and age. The correlation between MFI and NfL is important, as mitochondrial function is high correlative of neuronal dysfunction and neurodegeneration.

In the current study, plasma A/T/N biomarkers did not perform as well as the MFI biomarker for discrimination between ND and AD subjects. While prior studies have shown better AUC for pTau181 and Aβ ratios in plasma, this could be due to the size of our study or varying diagnostic criteria. Our blood-based MFI biomarker performed better than plasma pTau181, GFAP, NfL, and Aβ_42_/Aβ_40_ for discrimination between ND and AD subjects in this study and cohort. These data suggest that mitochondrial biomarkers could be used in combination with plasma A/T/N biomarkers to increase sensitivity and specificity in diagnostic and prognostic outcomes for AD. Future studies will address this possibility.

Our overall goal was to develop and test the utility of a blood-based mitochondrial biomarker that could be used as a response biomarker in future clinical trials. The MFI biomarker can detect mitochondrial dysfunction in human iPSC derived neurons treated with known mitochondrial toxins, iPSC derived neuron and astrocyte models of sporadic AD, 5xFAD mice, and human AD subjects. While the MFI biomarker did not discriminate between ND and MCI subjects, this is not surprising or concerning. Those with MCI often convert back to ND or eventually convert to AD so this reflects a heterogenous population with some mild cognitive impairment. The MFI did correlate with MMSE and CDR cognitive scores. Further studies are needed to interrogate the mechanisms of these correlations and understand if the MFI has any prognostic or predictive utility in cognition and AD.

We show that the MFI biomarker is relatively stable (∼30 hours in ACD tubes at room temperature) which allows for overnight shipments of blood to central labs if needed. This is not dissimilar to other labs that require fresh blood that are completed through commercialized central labs. However, the MFI biomarker requires additional clinical validation and development before implementation into clinical settings. A benefit of using a flow cytometry-based assay is it lends well to development of standalone high throughput instrumentation to measure the MFI; but this requires further engineering and cost/risk analyses.

Other considerations and questions include if a systemic mitochondrial biomarker reflects brain metabolism. Studies of AD reflect systemic mitochondrial dysfunction observed in blood cells (platelets, PBMCs), skin cells, and muscle; as such it’s not clear if a blood-based mitochondrial biomarker is required to reflect brain metabolism for utility. Improving systemic mitochondrial function is likely to have beneficial effects on brain outcomes, but this is to be determined. To attempt to answer the question of relationships between blood based mitochondrial biomarkers and brain metabolism, we have a current ongoing study examining the MFI biomarker, brain FDG-PET, brain MRI structure/volume, and newly FDA approved plasma A/T/N biomarkers (ptau217 and ptau217/Aβ_42_) within the same subjects.

With further clinical validation and intra-lab standard operating procedures, we believe the MFI and other potential blood based mitochondrial biomarkers could be leveraged in therapeutic clinical trails as response outcomes. This is especially critical for AD and related dementias given the increasing interest in therapies targeting metabolism. Its currently not feasible to know if a therapy fails in a clinical trial because it fails to engage its intended target (mitochondria/metabolism) or if it fails because the intended target is not disease modifying. The only way to overcome this is through response biomarker development specific to therapeutic targets, like mitochondrial function.

## Data Availability

All data produced in the present study are available upon reasonable request to the authors

## Acknowledgements and Funding

Research reported in this publication was supported by the National Center For Advancing Translational Sciences of the National Institutes of Health under Award Number UL1TR002366, the Margaret “Peg” McLaughlin and Lydia A. Walker Opportunity Fund, the University of Kansas, and Alzheimer’s Disease Center P30AG072973. We acknowledge the Flow Cytometry Core Laboratory, which is sponsored, in part, by the NIH/NIGMS COBRE grant P30GM103326 and the NIH/NCI Cancer Center grant P30CA168524. The content is solely the responsibility of the authors and does not necessarily represent the official views of the National Institutes of Health.

## References

1. Jack CR, Jr., Barrio JR and Kepe V. Cerebral amyloid PET imaging in Alzheimer’s disease. Acta Neuropathol 2013; 126: 643–657.

2. Flannagan K SJ, Hauger BM,, Troutwine BR LC, Strope TA,, Csikos Drummond V GC, et al. Cell type and sex specific mitochondrial phenotypes in iPSC derived models of Alzheimer’s disease. Front Mol Neurosci 2023; 16.

3. Troutwine BR, Strope TA, Franczak E, et al. Mitochondrial function and Aβ in Alzheimer’s disease postmortem brain. Neurobiol Dis 2022; 171: 105781.

4. Weidling IW, Wilkins HM, Koppel SJ, et al. Mitochondrial DNA Manipulations Affect Tau Oligomerization. J Alzheimers Dis 2020; 77: 149–163.

5. Wilkins HM, Troutwine BR, Menta BW, et al. Mitochondrial Membrane Potential Influences Amyloid-beta Protein Precursor Localization and Amyloid-beta Secretion. J Alzheimers Dis 2022; 85: 381–394.

6. Fukui H, Diaz F, Garcia S, et al. Cytochrome c oxidase deficiency in neurons decreases both oxidative stress and amyloid formation in a mouse model of Alzheimer’s disease. Proc Natl Acad Sci U S A 2007; 104: 14163–14168.

7. Pinto M, Pickrell AM, Fukui H, et al. Mitochondrial DNA damage in a mouse model of Alzheimer’s disease decreases amyloid beta plaque formation. Neurobiol Aging 2013; 34: 2399–2407.

8. Herholz K, Salmon E, Perani D, et al. Discrimination between Alzheimer dementia and controls by automated analysis of multicenter FDG PET. Neuroimage 2002; 17: 302–316.

9. Marcus C, Mena E and Subramaniam RM. Brain PET in the diagnosis of Alzheimer’s disease. Clin Nucl Med 2014; 39: e413–422; quiz e423-416.

10. Mosconi L, Berti V, Glodzik L, et al. Pre-clinical detection of Alzheimer’s disease using FDG-PET, with or without amyloid imaging. J Alzheimers Dis 2010; 20: 843–854.

11. Suppiah S, Didier MA and Vinjamuri S. The Who, When, Why, and How of PET Amyloid Imaging in Management of Alzheimer’s Disease-Review of Literature and Interesting Images. Diagnostics (Basel) 2019; 9.

12. Cardoso SM, Proenca MT, Santos S, et al. Cytochrome c oxidase is decreased in Alzheimer’s disease platelets. Neurobiol Aging 2004; 25: 105–110.

13. Cardoso SM, Santana I, Swerdlow RH, et al. Mitochondria dysfunction of Alzheimer’s disease cybrids enhances Abeta toxicity. J Neurochem 2004; 89: 1417–1426.

14. Chakravorty A, Jetto CT and Manjithaya R. Dysfunctional Mitochondria and Mitophagy as Drivers of Alzheimer’s Disease Pathogenesis. Front Aging Neurosci 2019; 11: 311.

15. Fisar Z, Hroudova J, Hansikova H, et al. Mitochondrial Respiration in the Platelets of Patients with Alzheimer’s Disease. Curr Alzheimer Res 2016; 13: 930–941.

16. Guo L, Tian J and Du H. Mitochondrial Dysfunction and Synaptic Transmission Failure in Alzheimer’s Disease. J Alzheimers Dis 2017; 57: 1071–1086.

17. J BS. Mitochondria in Alzheimer’s disease: An Electron Microscopy Study Alzheimer’s Disease & Treatment 2019.

18. Kish SJ, Bergeron C, Rajput A, et al. Brain cytochrome oxidase in Alzheimer’s disease. J Neurochem 1992; 59: 776–779.

19. Morris JK, Honea RA, Vidoni ED, et al. Is Alzheimer’s disease a systemic disease? Biochim Biophys Acta 2014; 1842: 1340–1349.

20. Parker WD, Jr. Cytochrome oxidase deficiency in Alzheimer’s disease. Ann N Y Acad Sci 1991; 640: 59–64.

21. Swerdlow RH. Mitochondria and Mitochondrial Cascades in Alzheimer’s Disease. J Alzheimers Dis 2018; 62: 1403–1416.

22. Wightman DP, Jansen IE, Savage JE, et al. A genome-wide association study with 1,126,563 individuals identifies new risk loci for Alzheimer’s disease. Nat Genet 2021; 53: 1276–1282.

23. Harwood JC, Leonenko G, Sims R, et al. Defining functional variants associated with Alzheimer’s disease in the induced immune response. Brain Commun 2021; 3: fcab083.

24. Lakatos A, Derbeneva O, Younes D, et al. Association between mitochondrial DNA variations and Alzheimer’s disease in the ADNI cohort. Neurobiol Aging 2010; 31: 1355–1363.

25. Swerdlow RH, Hui D, Chalise P, et al. Exploratory analysis of mtDNA haplogroups in two Alzheimer’s longitudinal cohorts. Alzheimers Dement 2020; 16: 1164–1172.

26. Andrews SJ, Fulton-Howard B, Patterson C, et al. Mitonuclear interactions influence Alzheimer’s disease risk. Neurobiol Aging 2020; 87: 138 e137–138 e114.

27. Carrieri G, Bonafe M, De Luca M, et al. Mitochondrial DNA haplogroups and APOE4 allele are non-independent variables in sporadic Alzheimer’s disease. Hum Genet 2001; 108: 194–198.

28. Wilkins HM and Swerdlow RH. Mitochondrial links between brain aging and Alzheimer’s disease. Transl Neurodegener 2021; 10: 33.

29. Jawdat O, Rucker J, Nakano T, et al. Resistance exercise in early-stage ALS patients, ALSFRS-R, Sickness Impact Profile ALS-19, and muscle transcriptome: a pilot study. Sci Rep 2024; 14: 21729.

30. Macchi Z, Wang Y, Moore D, et al. A multi-center screening trial of rasagiline in patients with amyotrophic lateral sclerosis: Possible mitochondrial biomarker target engagement. Amyotroph Lateral Scler Frontotemporal Degener 2015; 16: 345–352.

31. Statland JM, Moore D, Wang Y, et al. Rasagiline for amyotrophic lateral sclerosis: A randomized, controlled trial. Muscle Nerve 2019; 59: 201–207.

32. Taylor MK, Burns JM, Choi IY, et al. Protocol for a single-arm, pilot trial of creatine monohydrate supplementation in patients with Alzheimer’s disease. Pilot Feasibility Stud 2024; 10: 42.

33. Vidoni ED, Choi IY, Lee P, et al. Safety and target engagement profile of two oxaloacetate doses in Alzheimer’s patients. Alzheimers Dement 2021; 17: 7–17.

34. Wilkins HM, Koppel SJ, Bothwell R, et al. Platelet cytochrome oxidase and citrate synthase activities in APOE epsilon4 carrier and non-carrier Alzheimer’s disease patients. Redox Biol 2017; 12: 828–832.

35. Wilkins HM, Mahnken JD, Welch P, et al. A Mitochondrial Biomarker-Based Study of S-Equol in Alzheimer’s Disease Subjects: Results of a Single-Arm, Pilot Trial. J Alzheimers Dis 2017; 59: 291–300.

36. Committee NS. Coding Guidebook for the NACC Neuropathology Data Form. 11 ed. 2020.

37. Muratore CR, Srikanth P, Callahan DG, et al. Comparison and optimization of hiPSC forebrain cortical differentiation protocols. PLoS One 2014; 9: e105807.

38. Muratore CR, Rice HC, Srikanth P, et al. The familial Alzheimer’s disease APPV717I mutation alters APP processing and Tau expression in iPSC-derived neurons. Hum Mol Genet 2014; 23: 3523–3536.

39. Shaltouki A, Peng J, Liu Q, et al. Efficient generation of astrocytes from human pluripotent stem cells in defined conditions. Stem Cells 2013; 31: 941–952.

40. Chatterjee P, Pedrini S, Stoops E, et al. Plasma glial fibrillary acidic protein is elevated in cognitively normal older adults at risk of Alzheimer’s disease. Transl Psychiatry 2021; 11: 27.

41. Cicognola C, Janelidze S, Hertze J, et al. Plasma glial fibrillary acidic protein detects Alzheimer pathology and predicts future conversion to Alzheimer dementia in patients with mild cognitive impairment. Alzheimers Res Ther 2021; 13: 68.

42. Giacomucci G, Mazzeo S, Bagnoli S, et al. Plasma neurofilament light chain as a biomarker of Alzheimer’s disease in Subjective Cognitive Decline and Mild Cognitive Impairment. J Neurol 2022; 269: 4270–4280.

43. Mazzeo S, Ingannato A, Giacomucci G, et al. Plasma neurofilament light chain predicts Alzheimer’s disease in patients with subjective cognitive decline and mild cognitive impairment: A cross-sectional and longitudinal study. Eur J Neurol 2024; 31: e16089.

44. Tissot C, A LB, Therriault J, et al. Plasma pTau181 predicts cortical brain atrophy in aging and Alzheimer’s disease. Alzheimers Res Ther 2021; 13: 69.

45. Janelidze S, Mattsson N, Palmqvist S, et al. Plasma P-tau181 in Alzheimer’s disease: relationship to other biomarkers, differential diagnosis, neuropathology and longitudinal progression to Alzheimer’s dementia. Nat Med 2020; 26: 379–386.

46. Wojdala AL, Bellomo G, Toja A, et al. CSF and plasma Abeta42/40 across Alzheimer’s disease continuum: comparison of two ultrasensitive Simoa((R)) assays targeting distinct amyloid regions. Clin Chem Lab Med 2024; 62: 332–340.

